# The Effect of Selenium on Hashimoto’s Thyroiditis; Systemic Review and Metaanalysis

**DOI:** 10.1101/2020.04.15.20066001

**Authors:** Ehsan Alijani

## Abstract

**Background:** Selenium forms a significant component of seleno-proteins in the body. Seleno-methionine is integrated into proteins instead of methionine and acts as a storage pool. In proteins, the active form of Selenium is seleno-cysteine.in this review we aim to prevail the results of selenium effect on thyroid status in recent clinical trials. The systemic review aims to find out the correlation between supplementation of Selenium and anti-TPO antibodies and T4 levels in Hashimoto’s Thyroiditis. Selenium supplementation decreases the level of anti-TPO antibodies. The supplementation of Selenium increases the level of T4 levels.

**Methods:** The mean and standard deviation (SD) of all 8 studies was were calculated. One of the researches had all the information in figures, and only the levels of anti-TPO antibodies and FT4 were obtained. Heterogenicity was estimated using I2.

**Results:** The p-value calculated for Anti-TPO by SPSS of the eight groups had a p-value of 0.142. The p-value calculated for T4 levels by SPSS of the five groups had a p-value of 0.239. The heterogenicity test was zero after the I2 test. The studies that were included in the systematic review were assessed by Prisma diagram and selected among the articles resulted from search keywords selenium and anti tpo and hashimotho thyroiditis on different data sources. All the participants were evaluated by sex, ages, duration of the study and the levels of anti tpo ab and thyroxin (t4) and then using SPSS software to deploy metanalysis in the systematic review.

**Conclusion:** in 6 of 8 studies it was a relation of selenium and thyroid status, whereas 2 studies were not. As We ran meta analysis on the data we realized that there is not significant desired effect from selenium on thyroid antibodies against previous metanalysis done by other researchers.

## INTRODUCTION

Minerals are inorganic substances required by the human body for normal functioning. The body does not support synthesis or manufacture minerals; therefore, minerals are provided in the diet (Mason, 2012). Minerals are divided into two broad categories: macro-minerals and micro-minerals. All the metals are essential for the body, but micro-nutrients are required in a minute or trace amounts. Adults need more than 100mg daily and make less than 1 percent of total body weight. Adults should take between 1 to 100mg daily of trace minerals, and trace minerals make less than 0.01 percent of the whole-body weight. Ultra-trace minerals are required by the body in trace amounts lower than micro-minerals; their daily intake is less than one microgram (Kipp et al., 2015).

Selenium is one of the trace minerals required by the body. It has several roles in biological roles in the human body. The first selenium deficiency was identified in rats, which were devoid of vitamin E (Schwartz, 1957). The mice had a liver injury, which could be reversed by supplementing the mice’s diet with selenium. Toxicity is common with selenium supplementation attributed to the fact that it has a narrow optimum range. Any slight increase above the required limit is disastrous to the individual. Supplementation is necessary for patients or individuals who are deficient in the trace mineral only. People with a normal range of serum selenium are advised against taking selenium to avoid toxicity.

Minerals are not synthesized in the body because they are elements; therefore, they must be provided in the diet. Like any other mineral, selenium is obtained by eating plants or animals or from drinking water. Plants get minerals from the soil. Therefore, the selenium content in plants is influenced by the type of soil in which they are grown. Plant sources of selenium include brazil nut, cereals, and grains. Water contains very minute selenium concentrations (Barceloux DG, 1999). Animal-based sources of selenium are seafood, kidney, liver, and meat. Selenium is usually found in two forms: the organic forms are seleno-cysteine and seleno-methionine (Lavender OA, 1992); the inorganic forms are mostly used for supplementation. The organic forms of selenium are selenite and selenite (Holben DH, 1999).

The bioavailability of selenium is more than 50 percent. Seleno-methionine is actively absorbed in the small intestine by the methionine absorptive pathway (Thompson C, 1998). The absorptive of seleno-cysteine is not well known. Absorption of organic selenium in supplements passively takes place in the duodenum. Individual selenium status in plasma is uncorrelated, with its absorption making regulation difficult.

Selenium form a significant component of seleno-proteins in the body. Seleno-methionine is incorporated into proteins in place of methionine and serves as a storage pool. In proteins, the active form of selenium is seleno-cysteine. Seleno-cysteine can be synthesized from seleno-methionine, absorbed directly from the intestines, or synthesized y replacement of an oxygen residue on serine while it’s still to a specific tRNA. Selenium is released from the catabolism of seleno-methionine and seleno-cysteine. The selenium is excreted principally by the kidney in the urine.

More than 30 seleno-proteins have been identified. Only glutathione peroxidase and iodothyronine deiodinase are well understood. Glutathione peroxidase exists in four forms; it acts as an antioxidant defense while iodothyronine deiodinase exists in three types; it acts as a catalyst in thyroid hormone production. Other seleno-protein include selenoprotein P and seleno-protein synthase. Some seleno-proteins have well-defined roles while others their roles are not well understood and are under investigation (Holben, 1999).

Other potential roles of selenium have been hypothesized but yet well developed and are under study or investigation. Selenium plays are a role in enhancing the immune system. High concentrations of the mineral are found in tissues with hematopoietic and immune functions such as the liver, spleen, and lymph nodes. This suggests that selenium may have a function in the immune system. Studies have shown a linear relationship between selenium and the reduction in CD4 cells in HIV patients (Look MP, 1996). The depletion of selenium stores in tissues has been associated with impaired cell-mediated immunity. Natural killer cell activity is enhanced by selenium supplementation in individuals with selenium deficiency (Kiremidjian-Schumacher L, 1994). Selenium also decreases inflammatory activity if the thyroid in patients with autoimmune thyroiditis. It also reduces the risk of postpartum thyroiditis in women with antibodies against thyroid peroxidase. A relationship between selenium and cancer mortality has been developed, and studies are ongoing tor use of selenium as a prevention measure for cancer.

Selenium has also been hypothesized in the reduction of cardiovascular diseases. This is because glutathione peroxidase, a seleno-protein, reduces hydrogen peroxide and other molecules with oxidative potential, thus protecting lipid membranes, inhibiting modification of low-density protein and suppressing platelet aggregation. The effects of glutathione peroxidase suggest that supplementation with selenium would be protected against atherosclerosis.

Prospective and epidemiologic studies show mixed results, and further research is needed to prove the claim. Animal models suggest that low selenium levels improve glucose metabolism. Clinical studies in humans show no relationship between selenium and glucose metabolism and may increase the risk for type 2 diabetes.

Excessive intake of selenium leads to toxicity. The clinical manifestation of selenium toxicity includes nausea, emesis, diarrhea, hair loss, changes in nails, changes in mental status, loss of vision, and peripheral neuropathy. Brain imaging by magnetic resonance imaging show abnormalities that resemble reversible posterior encephalopathy syndrome. Toxicity occurs with excessive intake from the diet or excessive supplementation. In China, Enshi county chronic consumption of a plant-based diet with a selenium content of 5mh per day results in hair and nail loss, tooth decay, dermatologic lesions, and neurologic effects (Yang GQ, 1983).

In the united states, 201 experienced selenium toxicity after taking a liquid dietary supplement containing 200 times the labeled content of selenium. The selenium concentration ingested was more than 41,000 micrograms per day; this is almost 800 times the recommended dietary intake of 55 micrograms per day and 100 times the tolerable upper limit of 400 micrograms per day. In 1983, selenium poisoning occurred in 13 people who ingested mislabeled supplement.

Selenium in the diet is mainly from seafood, organ meats, and plant-based food; the concentration in the plant depends on the soil selenium concentrations. Selenium is a mineral and thus found in soil. The recommended dietary intake for selenium for young children is 20mcg per day and 55mcg per day for adults.

The deficiency of selenium has significant health implications. They include skeletal muscle dysfunction, cardiomyopathy, mood disorders, impaired immune function, macrocytosis, and white nailbeds. Keshan disease, due to selenium deficiency, is a cardiomyopathic disease affecting mainly children and women of childbearing age. Its more prevalent in China and has been linked to selenium deficiency. The geographic distribution of Keshan disease in China is attributed to the local diet, which is almost devoid of selenium. The disease positively responds to supplementation with selenium. (Ishida T, 2003). Selenium deficiency has been reported in patients receiving total parenteral nutrition. This is attributed to the fact that selenium is excluded in the trace elements added to parenteral nutrition. The patients present with cardiomyopathy and skeletal dysfunction. Selenium deficiency is described in a child with increased selenium loss in the chylous fluid due to lymphangiomatosis, despite being supplemented with selenium in the total parenteral nutrition.

Selenium is critical to the functioning of D1. These were demonstrated by a decrease in hepatic D1 activity of selenite deficient rats and that demonstration that D1 could be labeled with selenium. The efficiency of selenium deficiency on the synthesis of intracellular seleno-proteins, such as selenodeiodenidase, depends on the tissue under examination. For example, thyroidal D1 activity is preserved, while that in the liver declines sharply, serum T3 concentrations increase, and serum T4 concentration does not change. Selenium deficiency also decreases D1 activity in the kidney; this is accompanied by a decrease in D1 mRNA, which does not occur in the liver. Selenium deficiency also occurs in patients receiving diets restricted in their protein content, such as phenylketonuria, and also it is seen in elderly patients.

Patients with selenium deficiency, serum T4 concentrations, and the serum T4 to T3 are slightly increased while serum TSH concentrations are normal. In the endemic goiter region in Africa, selenium deficiency also is prevalent. When selenium was given to the iodine-deficient people, their thyroid function deteriorated, as seen by an increase in serum TSH concentrations and a decrease in the serum T3 concentrations, suggesting that the reduction in D1 activity during selenium deficiency can protect against iodine deficiency, by reducing inner ring deiodination of T4 and T3 (Lewis E, Bravermann, 2019).

A randomized control study in patients with mild but active Graves’ ophthalmopathy showed a significant response to selenium supplements of 200 micrograms per daily. The improvement was primarily due to a reduction in soft-tissue swelling and palpebral width. The quality of scores also improved significantly and was sustained twelve months after the initiation of treatment. No side effects were reported. A 6-month course of selenium appears to be a safe and effective treatment in mild active Graves’ ophthalmopathy.

Selenium administration in patients with Hashimoto’s thyroiditis reduces the level of anti-TPO antibodies. In a randomized control trial, administration of 200 micrograms of selenium daily to pregnant women, reduced the postpartum rise of anti-TPO antibodies and also decreased the incidence of PPT and hypothyroidism compared to control women with TPO antibodies in pregnancy who did not receive the drug.

Inadequate selenium supply is associated with the manifestation of various human diseases such as Keshan and Kashin-Beck disease, cancer, impairment of immune function, neurodegenerative diseases, age-related disorders, and disturbance of the thyroid hormone axis. Selenium deficiency, in conjunction with iodine deficiency, contributes to the pathogenesis of myxedematous cretinism. The recent identification of various distinct selenocysteine containing proteins, encoded by 25 human genes, provides information on the molecular and biochemical basis of beneficial and possible adverse effects of this trace or micromineral.

The thyroid gland is one of the organs in the human body that contains the highest concentration of selenium per mass unit compared to other endocrine organs and the brain. Seleno-proteins involved in cellular antioxidative defense systems and redox control, such as glutathione peroxidase and thioredoxin reductase family, are included in the protection of the thyroid gland from excess hydrogen peroxide and reactive oxygen species produced y follicles for the biosynthesis of thyroid hormones. The iodothyronine deiodinases (DIO 1,2,3), which are a seleno-proteins, are involved in the activation and deactivation of thyroid hormones. They are associated with developmental, cell, and pathology expression patterns. Nutritional supply of selenium usually is sufficient for adequate expression of functional DIO enzymes, except for long term parenteral nutrition and other diseases impairing gastrointestinal absorption of selenium compounds. The nutritional supply of selenium for protection of the thyroid gland and the synthesis of some of the abundant seleno-proteins of the glutathione peroxidase and TrxR family might limit their proper expression under pathophysiological conditions (Josef Köhrle,2009).

## AIMS/OBJECTIVE

The systemic review aims to find out the correlation between selenium supplementation and anti-TPO antibodies and T4 levels in Hashimoto’s Thyroiditis.

## HYPOTHESIS

Selenium supplementation decreases the level of anti-TPO antibodies. Selenium supplementation increases the level of T4 levels.

## MATERIAL AND METHOD

1. The question under study was well defined, and inclusion criteria determined. The inclusion criteria used were any article published from the year 2000 to 2019. The participants were adults above 18 years of age and are suffering from autoimmune thyroiditis.
2. A literature search was conducted using various human studies databases like Cochrane, Medline, Biomed, Google Scholar, Scopus, and Embase. Smaller journals that are not in the databases were also reviewed, and non-published articles from like abstracts presented in conferences and current research were also included.
3. The terms used for the search were selenium, autoimmune thyroiditis, and anti-TPO antibodies.
4. The articles were pulled, those that were not written in English were translated to English using ginger translate.
5. The quality of the articles that were selected was determined using the Jadad scoring scale, also known as the Oxford quality scoring system. Materials with a Jadad score of 3 and below were excluded from the list and not used for the meta-analysis.
6. The abstracts of the selected articles were read, the exclusion and inclusion criteria were used to narrow down into the required and relevant sources.
7. The whole articles were reviewed by multiple reviewers to minimize bias and those that didn’t meet the inclusion criteria removed. Eight reviews that met all the requirements were downloaded and data abstracted into a table form, as shown below.
8. After abstraction, data analysis was carried out. The mean and standard deviation (SD) were calculated. One of the researches had all the information in figures, and only the levels of FT4 and anti-TPO antibodies were obtained. Two random effect analysis was performed for the mean difference in the selected studies for FT4 and anti-TPO antibodies level. Heterogenicity was estimated using I2. If the heterogenicity index were less than 25 percent, the random effect would be fixed, and if it is higher than 25 percent, the random fact will be selected. In the comprehensive meta-analysis, paired mean difference and variance were chosen. The mean difference between the setpoint and after treatment in test groups and the sample size was also entered. The summation of before calculated the variation of mean deviation and after variances (SD)^2.

### Included studies

The studies that were included in the study were Gartner 2003 from Greece, which has a sample size of 70. All the participants were females, and the treatment group was supplemented, with 200 grams of selenium for six months. Turker 2006 from turkey. The randomized trial had a sample size of 88, and the control group was supplanted with 200 rams of selenomethionine for six months. Duntas 2003 from Greece with a sample size of 65 and the treatment group supplemented with selenomethionine for three months. Esposito 2016 from Spain with a sample size of 76 and supplemented with 166 grams of selenomethionine for six months. Farias 2015 from brazil with a sample size of 55, and the study lasted three months. Eske 2014 from holland with a sample size of 61, and the study was carried out for six months. Pirola 2016 from Spain with a sample size of 192 and the study lasted for four months and Kachouei 2018 from Iran with a sample size of 70, patients supplemented with sodium selenite for three months

## Results

The results obtained from the 8-literature reviewed were analyzed using SPSS software version 22 to come up with the data used to give a conclusion on the metanalysis. The research done by Gartner 2002 showed that anti-TPO antibodies decreased significantly in the treatment group supplemented with selenium to 63.5% and 88% in the control group given placebo. Eske et al., 2014 showed that there was no significant difference between the treatment and control or placebo group after selenium supplementation. Duntas 2003 showed a decrease in anti-TPO antibodies and TSH. Turker, 2006, the anti-TPO value was 4 IU/ml, and the TSH level was 0.025. Esposito 2016, showed no difference between the treatment and placebo groups. Farias 2015 showed a 5% decrease in anti-TPO antigens at three months and a 20% decrease at six months. Pirola study done in 2016, 17.2% of patients received euthyroidism after supplementation with selenium, and Kachouei 2018, the mean of anti-TPO antibodies was 682.18 before selenium supplementation and 522 after supplementation.

The p-value calculated for Anti-TPO by SPSS of the eight groups resulted in a p-value of 0.142. The p-value calculated for T4 levels by SPSS of the five groups resulted in a p-value of 0.239. The heterogenicity test was zero after the I2 test.

### Forest plots

Forest plots are used to show if a correlation exists between the selenium supplementation and anti-TPO antibodies and the T4 level. The plot is constructed using means and 95 percent confidence interval. A vertical line running through the zero marks show the point of no difference. All studies were plotted. The diamond shape overlaps the line of no difference in both anti-TPO and T4 levels, indicating that the meta-analysis shows no effect when patients with Hashimoto’s thyroiditis are supplemented with selenium on the level on anti-TPO antibodies and T4.

**Figure 1.**
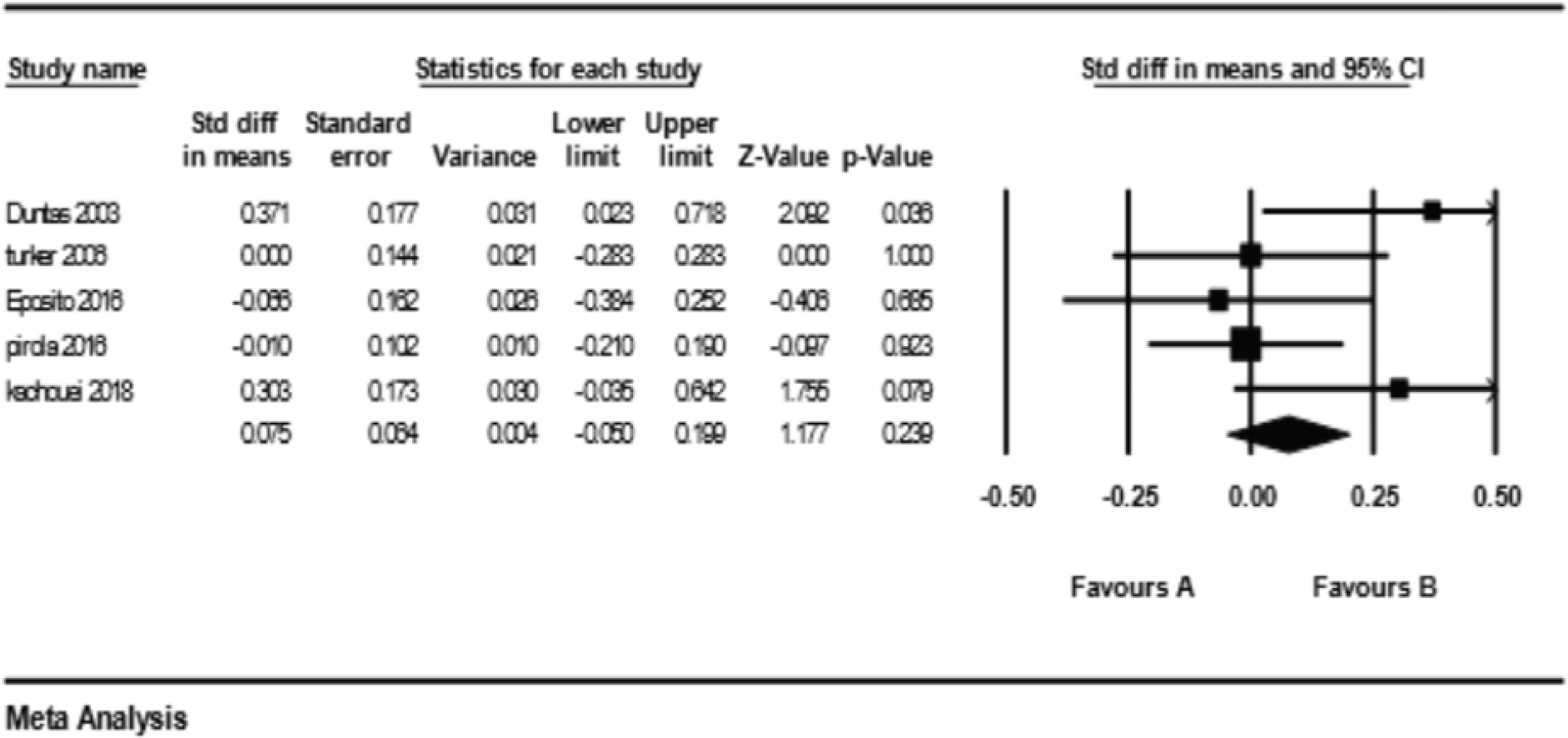
A plot of T4 level std mean difference before and after selenium supplementatio

**Figure 2.**
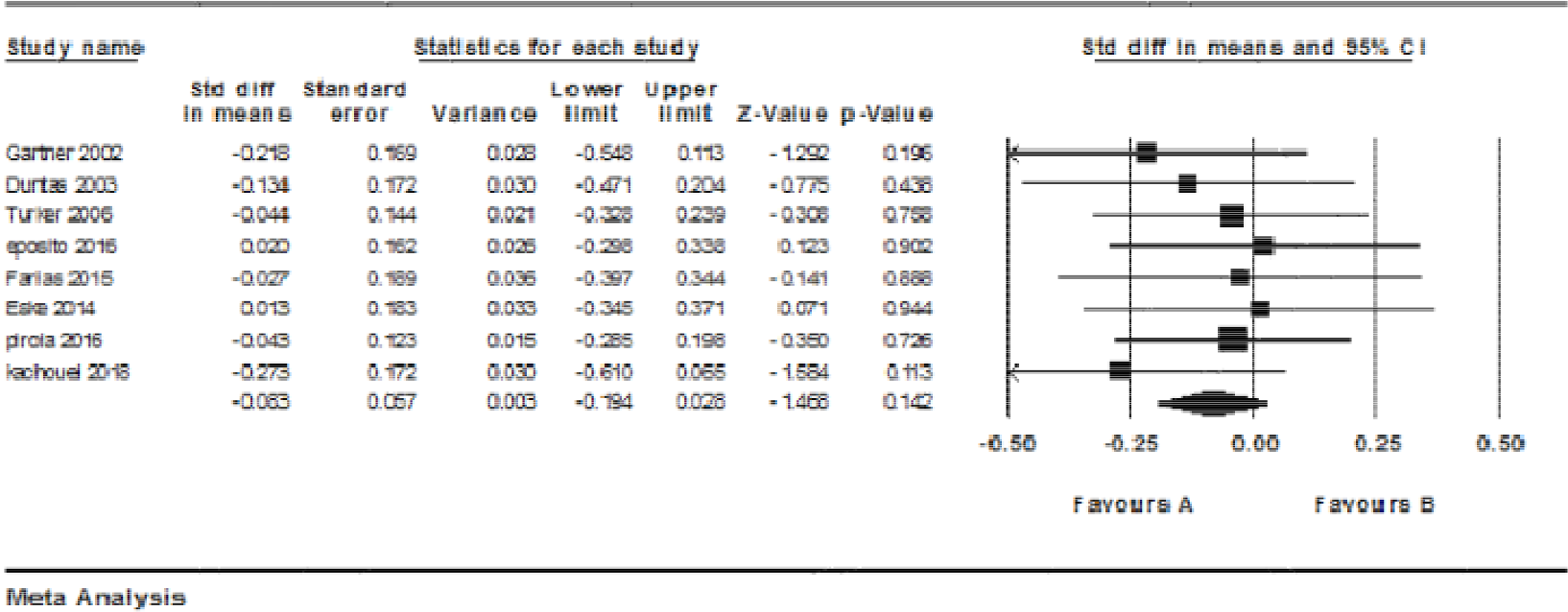
plot of Anti tpo ab level std mean difference before and after selenium supplementation

### Literature review and discussion

Gartner et al. 2002 performed a blinded, placebo-controlled prospective study in female patients (n 70; mean age, 47.5 - 0.7 years) with autoimmune thyroiditis and thyroid peroxidase antibodies (TPOAb) and Tg antibodies (TgAb) above 350 IU/ml. The aim of the experiment was to explore whether supplementation lowers thyroid peroxidase antibodies concentrations. The primary endpoint of the study was the change in TPOAb concentrations. Secondary endpoints were changes in TgAb, TSH, and free thyroid hormone levels as well as the ultrasound pattern of the thyroid and quality of life estimation. Patients were randomized into two age- and antibody (TPOAb)-matched groups; 36 patients received 200 g (2.53 mol) sodium selenite/d, orally, for 3months, and 34 patients received a placebo. All patients were substituted with L-T4 to maintain TSH within the normal range.The mean TPOAb concentration decreased significantly to 63.6% (P 0.013) in the selenium group vs. 88% (P 0.95) in the placebo group. A subgroup analysis of those patients with TPOAb greater than 1200 IU/ml revealed a mean 40% reduction in the selenium-treated patients compared with a 10% increase in TPOAb in the placebo group.

In conclusion, seleno-methionine supplementation had no positive effect on thyroid echogenicity or TPOAb in the patients. However, we observed a Seleno-methionine dependent downregulation of the IFN γ-inducible chemokines, especially CXCL-9 and -10, which serve as helpful biomarkers in the future selenium supplementation trials.

Manevskaa et all; 2019 conducted a randomized clinical trial. Five hundred thyroid patients, males, and females mean age 46 ± 19 years, with diagnosed HT, were included in the study. Euthyroid forms of HT were treated with Se only, while patients with thyroid-stimulating hormone (TSH) > 10 μIU/mL were treated with both substitutional therapy of levothyroxine and Selenium.

In around 37% of the patients treated with Se 3 × 50 μg/day with anti TPO> 1,000 IU/mL, anti TPO remained unchanged after 12 months, while 24.16% had anti TPO< 500 IU/mL and 38.20% had anti TPO between 500 and 1,000 IU/ml. Eighty-three out of 150 (55.33%) patients treated with Se 2 × 50 μg/day with anti-TPO between 500 and 1,000 IU/mL responded. More than half of the patients (91/172, 52.90%) with anti-TPO < 500 IU/mL treated with Se 50 μg/day normalized in 1 year. In a hypothyroid group of patients, 12 months after treatment with levothyroxine and Se, 47.18% were responders with anti-TPO> 1,000 IU/mL, while 79.20% with anti-TPO between 500 and 1,000 IU/mL. In the euthyroid group (Se only), the biggest response (30.56%) was seen in patients with the highest titer of anti-TPO> 1,000 IU/mL.

A total of 196 patients with autoimmune thyroiditis were recruited in the study by Pirola et al., 2006. The trial aims to find out whether selenium supplementation can restore euthyroidism in subclinical hypothyroid patients with autoimmune thyroidism. Patients were assigned to receive (case) or not receive (control) an oral seleno-methionine treatment. Cases received 83 mcg seleno-methionine/day orally for four months.Patients were assigned to receive (case) or not receive (control) an oral seleno-methionine treatment. Cases received 83 mcg seleno-methionine/day orally for four months. The results obtained showed that selenium supplementation could restore euthyroidism caused by Hashimoto’s thyroiditis.

Kachouei et al. 2019 performed a randomized clinical trial, 70 patients with autoimmune hypothyroidism. The aim was to determine the effect of selenium and levothyroxine on Anti tpo level and levothyroxine on its own. They randomly divided into two groups of 35 each; the first group was treated with oral selenium treatment with levothyroxine (LT4), and to the second group along with LT4, the placebo was also prescribed. Serum selenium level, thyroid hormones, and anti-thyroid hormone antibodies before and after three months of treatment in both groups, were determined, and the results were analyzed using SPSS software.

The mean of the serum anti-thyroid peroxidase serum level in the intervention group before and after treatment was 682.18 ± 87.25 and 522.96 ± 47.21 and the difference before and after treatment was statistically significant (P = 0.021). The level of this antibody before and after treatment in the control group was 441 ± 53.54 and 501.18 ± 77.68, and no significant differences between the two groups were observed before and after treatment (P = 0.42). They concluded that selenium could help reduce the levels of antibodies in patients with Hashimoto’s thyroiditis.

Pilli et al. 2015 performed a prospective, randomized, controlled study was conducted to evaluate the effect of two doses of seleno-methionine (Semet; 80 or 160 μg/day) versus placebo in euthyroid women with AIT, in terms of reduction of anti-thyroid antibodies, CXCL-9, -10 and -11 and improvement of thyroid echogenicity, over 12 months. Anti-thyroperoxidase antibody (TPOAb) levels remained unaffected by Semet Supplementation; anti-thyroglobulin antibody levels showed a significant reduction in the 160-Semet and the placebo group at 12 months.

Duntas et al., 2003 performed a randomized, placebo-controlled prospective study to investigate the effects of Se treatment on patients with autoimmune thyroiditis (AIT). Sixty-five patients aged 22–61 years (median age 48 years) with AIT were recruited into two groups. Group I (Gr I) 34 person was treated with seleno-methionine (Semet)200 mg, plus L-thyroxine (LT4) to maintain TSH levels between 0.3–2.0 mU/l, whereas group II (Gr-II) 31 received LT4 plus placebo for six months. There were no significant changes in antibodies against thyroglobulin levels between the groups. At the end of this study, Se levels were found to be statistically significantly increased in Gr I. serum levels of selenium increased substantially after oral administration, indicating excellent absorption of the supplement. Patients treated with selenium and levothyroxine showed a significant reduction in anti-TPO antibodies without affecting the metabolism of thyroid hormones.

Turker et al., 2006 carried out a study to investigate the long-term (9 months) effects of variable doses (200/100 mg/day) of L-seleno-methionine on autoimmune thyroiditis (AIT) and the parameters affecting the success rate of this therapy. The study was designed in three steps: (1) 88 female patients with AIT (mean age (40.1+-13.3 years) were randomized into two groups according to their initial serum TSH, thyroid peroxidase antibody (TPOAb) concentrations, and age. All the patients were receiving L-thyroxine to keep serum TSH>2 mIU/l.

Group S2 (n=48, mean TPOAb=803.9_+483.8 IU/ml) received 200 mg L-seleno-methionine per day, orally for 3 months, and group C (n=40, mean TPOAb=770.3 _+ 406.2 IU/ml) received placebo. A significant decrease in thyroglobulin antibody titers was only noted in group S2 (5.2%, P=0.01). L-seleno-methionine substitution suppresses serum concentrations of TPOAb in patients with AIT, but suppression requires doses higher than 100 mg/day, which is sufficient to maximize glutathione peroxidase activities. The suppression rate decreases with time. It was concluded that oral administration of 200 micrograms of L-seleno-methionine decreases serum anti-TPO antibodies.

Esposito,2016 conducted a placebo-controlled randomized prospective study; they enrolled untreated euthyroid patients with HT. The study aimed to determine the short time effects of L-seleno-methionine on the function of the thyroid gland in patients with Hashimoto’s Disease. Seventy-six patients were randomly assigned to receive l-selenomethionine 166 μg/day (SE n = 38) or placebo (controls n = 38) for 6 months. TSH, free T4 (FT4), free T3(FT3), TPOAb, and CXCL10 serum levels were assayed at time 0, after 3 and 6 months. FT4 levels were significantly decreased (P < 0.03) after three months, while FT3 increased (P < 0.04) after 3 and 6 months versus baseline values. In the control group, the FT3 decreased after 3 and 6 months (P < 0.02) compared to baseline. The results suggest the ineffectiveness of short-term L-seleno-methionine supplementation in HT.

Eskes et al.,2014 performed a randomized, placebo-controlled, double-blinded to evaluate in euthyroid TPO-Ab-positive women without thyroid medication, whether selenite decreases TPO antibodies and improved in the quality of life. Euthyroid (TSH 0.5–5.0 mU/l, FT4 10–23 pm) women with TPO-Ab ≥ 100 kU/l were randomized to receive 200 mcg sodium selenite daily (n = 30) or placebo (n = 31) for 6 months. TSH, FT4, TPO-Ab, selenium (Se), seleno-protein P (Sepp), and QoL were measured at baseline, 3, 6, and 9 months.

The conclusion made is, six-month selenite supplementation increases markers for selenium but does not have an effect on serum anti-TPO antibodies, TSK, or the quality of life between the treatment and control groups.

From the literature reviewed, there have been mixed results, some researchers argue that selenium supplementation increase anti-TPO antibodies, others suggest that selenium supplementation reduces the level of anti-TPO antibodies and others suggest no correlation at all.

## CONCLUSION

From the literature reviewed in the meta-analysis, six of the randomized clinical trials showed that selenium supplementation lowers the serum levels of anti-TPO antibodies while two experiments by Eskes et al., and Pirola et al. show that selenium supplementation does not affect anti-TPO antibodies. Despite various researches concluding that selenium supplementation significantly lower anti-TPO antibodies, my meta-analysis shows no significant statistical results in support of selenium supplementation lowering anti-TPO antibodies and FT4 Levels. My meta-analysis had a P-value of 0.142; the is more than 0.05; therefore, we reject the alternative hypothesis that selenium decreases anti-TPO antibodies. Selenium does not affect anti-TPO antibodies and T4.

Duntas et al. determined the rate of absorption of selenium. After oral administration of the serum levels of Se, increase drastically, indicating rapid absorption. Further studies should be performed to assess the bioavailability of Se. Intake alone cannot be conclusive about the effect of Se. The mechanism of action of Se on the immune system is not well established, and further studies need to be conducted on the same. Selenium biomarkers such as chemokines CXCL 9 and 10 induced y IFN y should be further investigated to help in the control of selenium supplementation. Further studies should be conducted to determine the modification of selenium supplements to make them useful for the treatment and prevention of Hashimoto’s Disease.

## Data Availability

The data is on the last page of file dhown in a table.

## APPENDIX

### Appendix A

**Prisma 2009 flow diagram**

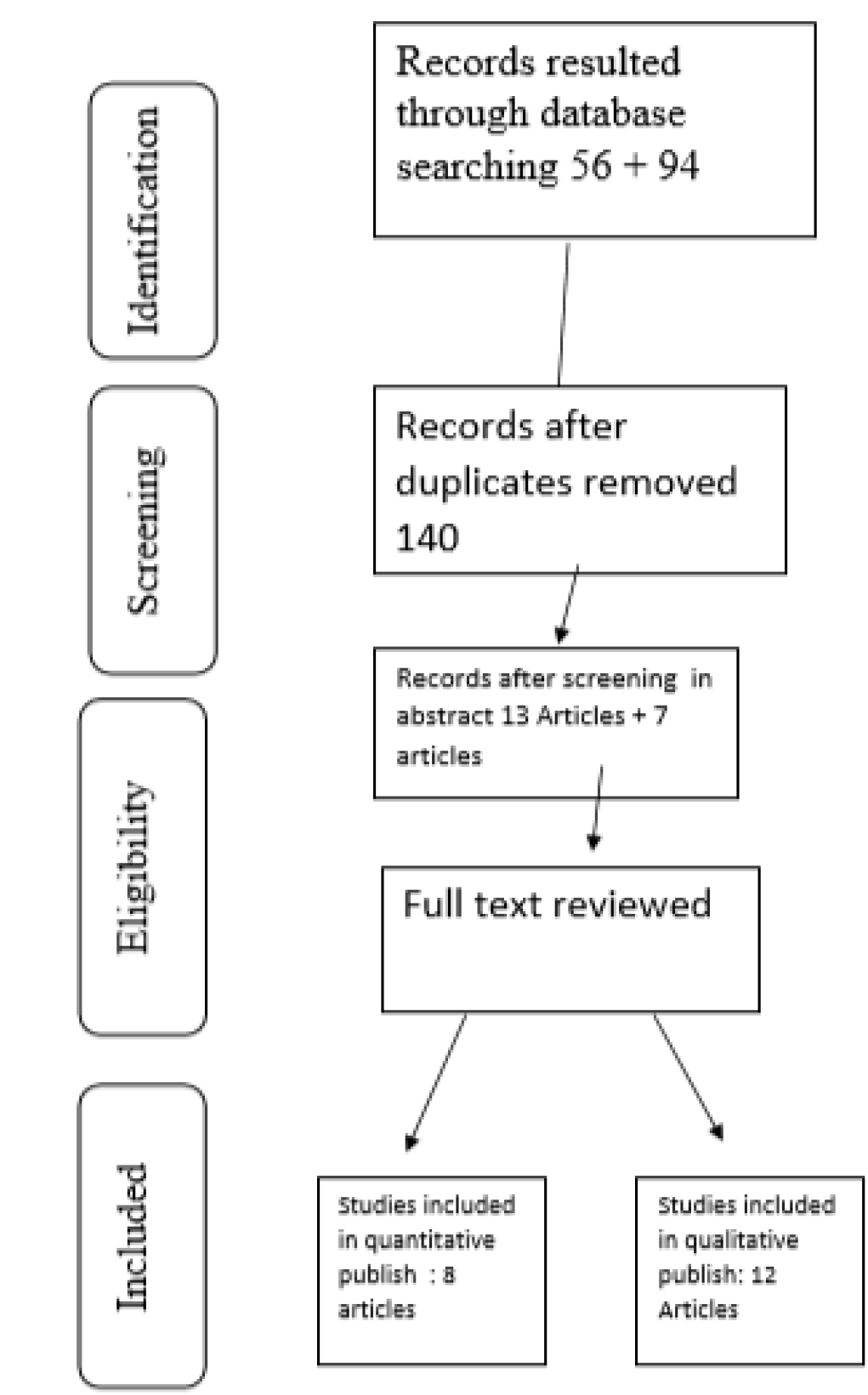
PRISMA 2009 FLOW diagram

### Appendix B

Data analyzed to plot the forest plot

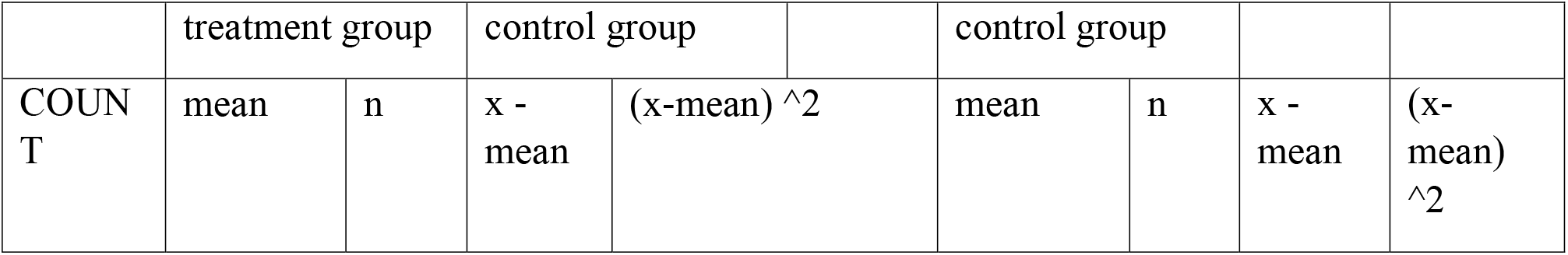
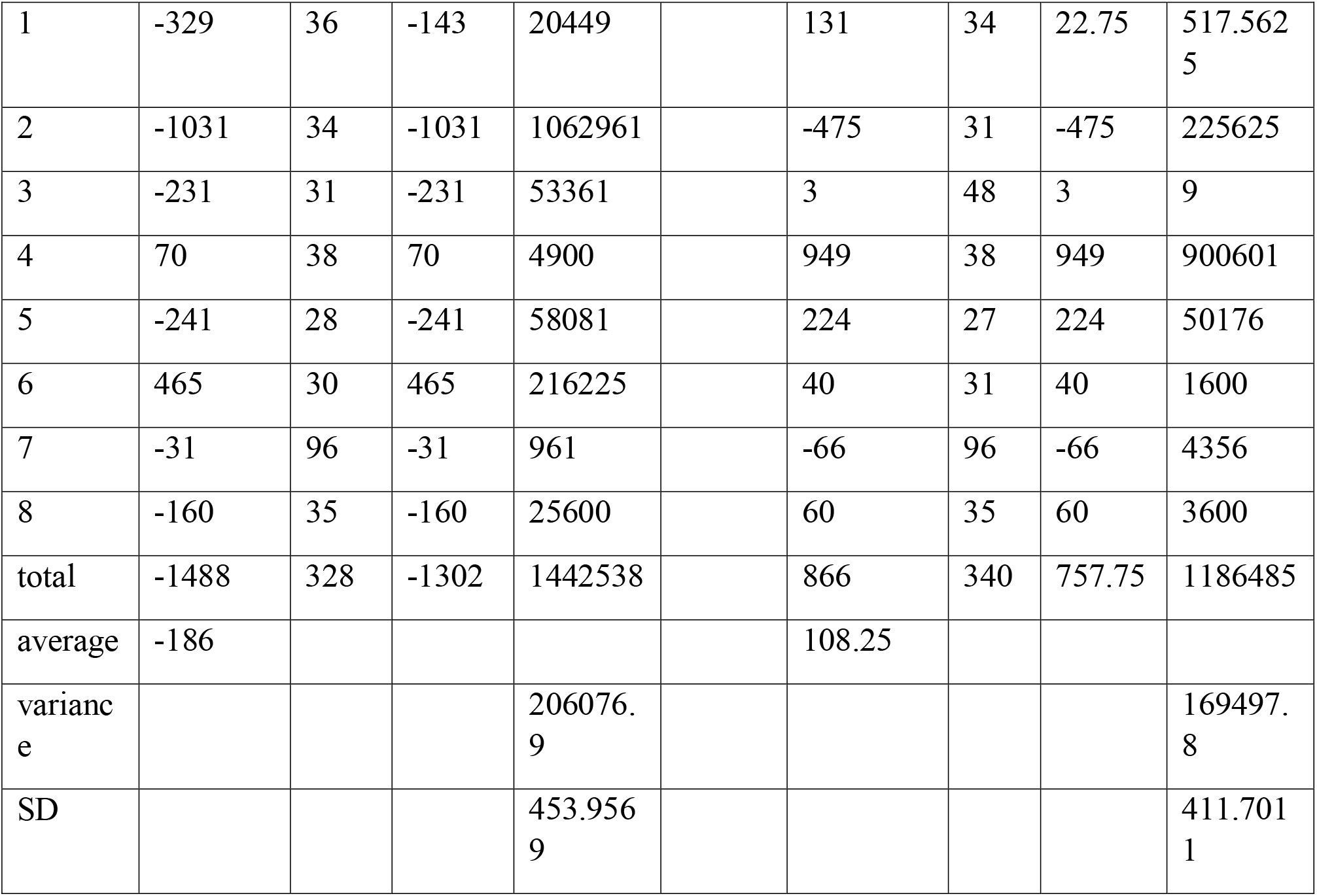

|  | treatment group |  | control group |  | control group |  |  |  |
| --- | --- | --- | --- | --- | --- | --- | --- | --- |
| COUNT | mean | n | x - mean | $(x - \text{mean})^2$ | mean | n | x - mean | $(x - \text{mean})^2$ |

|  |  |  |  |  |  |  |  |  |  |
| --- | --- | --- | --- | --- | --- | --- | --- | --- | --- |
| 1 | -329 | 36 | -143 | 20449 |  | 131 | 34 | 22.75 | 517.5625 |
| 2 | -1031 | 34 | -1031 | 1062961 |  | -475 | 31 | -475 | 225625 |
| 3 | -231 | 31 | -231 | 53361 |  | 3 | 48 | 3 | 9 |
| 4 | 70 | 38 | 70 | 4900 |  | 949 | 38 | 949 | 900601 |
| 5 | -241 | 28 | -241 | 58081 |  | 224 | 27 | 224 | 50176 |
| 6 | 465 | 30 | 465 | 216225 |  | 40 | 31 | 40 | 1600 |
| 7 | -31 | 96 | -31 | 961 |  | -66 | 96 | -66 | 4356 |
| 8 | -160 | 35 | -160 | 25600 |  | 60 | 35 | 60 | 3600 |
| total | -1488 | 328 | -1302 | 1442538 |  | 866 | 340 | 757.75 | 1186485 |
| average | -186 |  |  |  |  | 108.25 |  |  |  |
| variance |  |  |  | 206076.9 |  |  |  |  | 169497.8 |
| SD |  |  |  | 453.9569 |  |  |  |  | 411.7011 |

### Appendix C

**Tables1:** The characteristics of the trials include in the systematic review

Appendix C Tables 1: The characteristics of the trials included in the systematic review
| Study | Country | Duration(months) | Sample size | Intervention groups | LT4 treated | Age (years) | Female % |
| --- | --- | --- | --- | --- | --- | --- | --- |
| Gartner2002 | Germany | 6 | 70 | NaSe200<br>µg/day | All |  | 100% |
| Duntas2003 | Greece | 6 | 65 | Seme 200<br>µg/day | All |  | 86% |
| Turker2006 | Turkey | 3 | 88 | Seme200<br>µg/day | All |  | 100% |
| Eposito 2016 | spain | 6 | 76 | Seme 166<br>µg/day | None |  | 100% |
| Farias 2015 | Brazil | 3 | 55 | Seme200<br>µg/day | Some |  | 91% |
| Eske 2014 | Holland | 6 | 61 |  | None |  | 90% |
| Pirola 2016 | Spain | 4 | 192 | Seme(83<br>mcg) | None |  | 64% |
| Kachouei 2018 | Iran | 3 | 70 | SeNa(200<br>µg/day) | All |  | 64% |

